# NutriSighT: Interpretable Transformer Model for Dynamic Prediction of Hypocaloric Enteral Nutrition in Mechanically Ventilated Patients

**DOI:** 10.1101/2025.01.06.25320067

**Authors:** Mateen Jangda, Jayshil Patel, Jaskirat Gill, Paul McCarthy, Jacob Desman, Rohit Gupta, Dhruv Patel, Nidhi Kavi, Shruti Bakare, Eyal Klang, Robert Freeman, Anthony Manasia, John Oropello, Lili Chan, Mayte Suarez-Farinas, Alexander W Charney, Roopa Kohli-Seth, Girish N Nadkarni, Ankit Sakhuja

## Abstract

Achieving adequate enteral nutrition among mechanically ventilated patients is challenging, yet critical. We developed NutriSighT, a transformer model using learnable positional coding to predict which patients would achieve hypocaloric nutrition between days 3-7 of mechanical ventilation. Using retrospective data from two large ICU databases (3,284 patients from AmsterdamUMCdb – development set, and 6,456 from MIMIC-IV – external validation set), we included adult patients intubated for at least 72 hours. NutriSighT achieved AUROC of 0.81 (95% CI: 0.81 – 0.82) and an AUPRC of 0.70 (95% CI: 0.70 – 0.72) on internal test set. External validation with MIMIC-IV data yielded a AUROC of 0.76 (95% CI: 0.75 – 0.76) and an AUPRC of (95% CI: 0.69 – 0.70). At a threshold of 0.5, the model achieved a 75.16% sensitivity, 60.57% specificity, 58.30% positive predictive value, and 76.88% negative predictive value. This approach may help clinicians personalize nutritional therapy among critically ill patients, improving patient outcomes.

## INTRODUCTION

Optimal enteral nutrition (EN) is vital for critically ill patients requiring mechanical ventilation to meet their metabolic needs while mitigating complications [1,2]. Critical care guidelines recommend initiating early enteral nutrition in critically ill patients, but there is heterogeneity in the recommended caloric targets for the first week of intensive care unit (ICU) stay. For example, the European Society of Parenteral and Enteral Nutrition (ESPEN) advocates for hypocaloric nutrition (receipt of less than 70% daily caloric requirements) during the first week of ICU stay, but the American Society of Parenteral and Enteral Nutrition (ASPEN) recommends a broader caloric intake range of 12 to 25 kcal/kg, encompassing both hypocaloric and isocaloric nutrition strategies [3,4]. This variation reflects the dynamic nature of critical illness and the challenges in determining optimal nutrition within this highly heterogenous patient population [5–8]. Delivering adequate nutrition in this population is further complicated by challenges such as gastrointestinal dysfunction, hemodynamic instability, and frequent interruptions for procedures [9–11].

The first week of critical illness is divided into two distinct phases: the early acute period and the late acute period [1,12]. The early acute period spans the 48 hours of critical illness and is marked by hemodynamic instability and acute illness response. The late acute period, spanning days 3-7, is characterized by muscle wasting and evolving nutritional needs. Common clinical practice is to start a form of restrictive dose EN, such as trophic dose feeding, during the first 48 hours and progressively increase nutritional support during the late acute period to meet the evolving metabolic demands of critically ill patients.

However, these strategies are not personalized, and highly individualized nature of critical illness highlights a pressing need for tools that can dynamically identify patients likely to receive specific nutrition regimens. Addressing these challenges requires innovative methods integrating diverse clinical and temporal data to adapt nutritional interventions effectively.

Recent advancements in artificial intelligence (AI) can address these challenges [13,14]. Transformer models, known for their ability to model sequential data are powerful tools for analyzing clinical time series data [14,15]. The incorporation of learnable positional encoding [14,15], enhances the model’s ability to understand temporal relationships. Unlike fixed positional encodings, learnable encodings enable the model to optimally represent temporal dynamics and address complex clinical scenarios such as the predicting which patients are at risk of receiving hypocaloric nutrition [14,15].

In this study, we developed and externally validated an interpretable, transformer model, **NutriSighT**, to dynamically identify critically ill patients requiring mechanical ventilation who are at risk of receiving hypocaloric enteral nutrition during the late acute period of their critical illness. By focusing on this aspect, NutriSighT aims to address key gaps in nutritional management by offering actionable insights into patient-specific needs.

## RESULTS

### Patient Characteristics

A total of 3,284 patients from the AmsterdamUMCdb and 6,456 patients from the MIMIC-IV databases met the inclusion/exclusion criteria [16–18]. As shown in Table 1, AmsterdamUMCdb cohort had a higher proportion of younger patients (11.15% vs 9.15% in 18-39 year; p<0.001), males (64.58% vs 58.78%; p<0.001) and a lower BMI (median 24.8 kg/m² vs 28.1 kg/m²; p<0.001). Additional characteristics are provided in Supplementary Table 1.

**Table 1.**
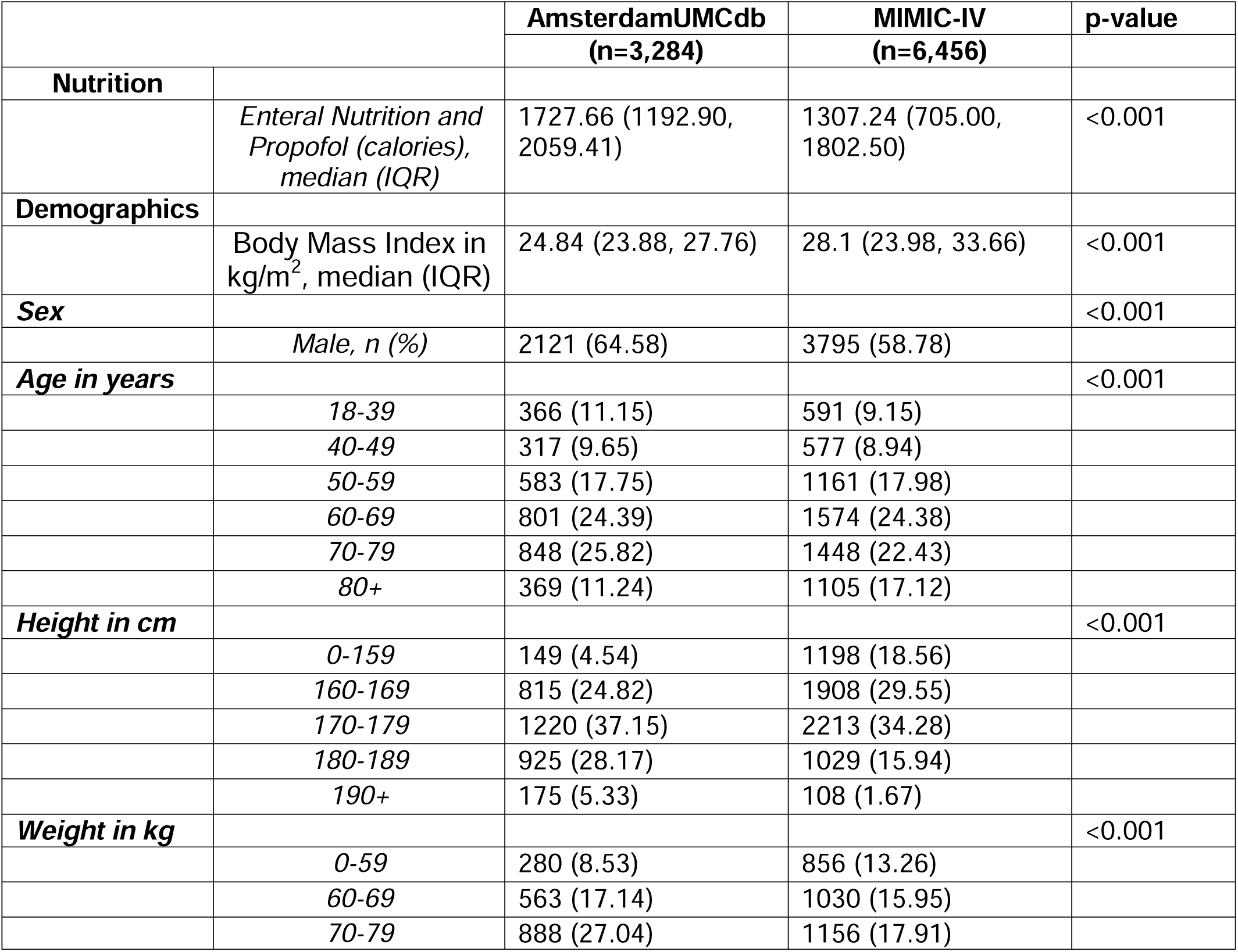
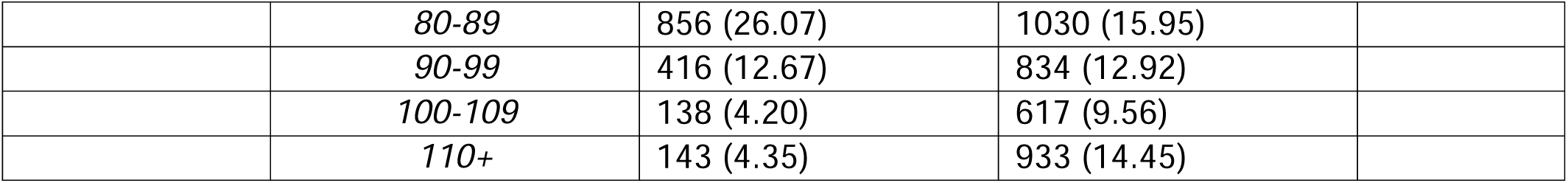
Patient Characteristics

|  |  | **AmsterdamUMCdb**<br>**(n=3,284)** | **MIMIC-IV**<br>**(n=6,456)** | **p-value** |
| --- | --- | --- | --- | --- |
| **Nutrition** |  |  |  |  |
|  | *Enteral Nutrition and Propofol (calories), median (IQR)* | 1727.66 (1192.90, 2059.41) | 1307.24 (705.00, 1802.50) | <0.001 |
| **Demographics** |  |  |  |  |
|  | Body Mass Index in kg/m<sup>2</sup>, median (IQR) | 24.84 (23.88, 27.76) | 28.1 (23.98, 33.66) | <0.001 |
| **Sex** |  |  |  | <0.001 |
|  | *Male, n (%)* | 2121 (64.58) | 3795 (58.78) |  |
| **Age in years** |  |  |  | <0.001 |
|  | 18-39 | 366 (11.15) | 591 (9.15) |  |
|  | 40-49 | 317 (9.65) | 577 (8.94) |  |
|  | 50-59 | 583 (17.75) | 1161 (17.98) |  |
|  | 60-69 | 801 (24.39) | 1574 (24.38) |  |
|  | 70-79 | 848 (25.82) | 1448 (22.43) |  |
|  | 80+ | 369 (11.24) | 1105 (17.12) |  |
| **Height in cm** |  |  |  | <0.001 |
|  | 0-159 | 149 (4.54) | 1198 (18.56) |  |
|  | 160-169 | 815 (24.82) | 1908 (29.55) |  |
|  | 170-179 | 1220 (37.15) | 2213 (34.28) |  |
|  | 180-189 | 925 (28.17) | 1029 (15.94) |  |
|  | 190+ | 175 (5.33) | 108 (1.67) |  |
| **Weight in kg** |  |  |  | <0.001 |
|  | 0-59 | 280 (8.53) | 856 (13.26) |  |
|  | 60-69 | 563 (17.14) | 1030 (15.95) |  |
|  | 70-79 | 888 (27.04) | 1156 (17.91) |  |
|  | *80-89* | 856 (26.07) | 1030 (15.95) |  |
|  | *90-99* | 416 (12.67) | 834 (12.92) |  |
|  | *100-109* | 138 (4.20) | 617 (9.56) |  |
|  | *110+* | 143 (4.35) | 933 (14.45) |  |

### Enteral Nutrition

Overall daily enteral nutrition support differed significantly between the AmsterdamUMCdb and MIMIC-IV cohorts. Patients in the AmsterdamUMCdb dataset received a median daily EN intake of 1440.0 mL (IQR: 1000.40–1723.80), compared to 756.48 mL (IQR: 228.72–1199.97) in MIMIC-IV (p<0.001). This difference was also evident in the daily caloric intake derived from EN, which was notably higher in AmsterdamUMCdb (median: 1702.99 kcal, IQR: 1155.26–2039.66) relative to MIMIC-IV (median: 989.20 kcal, IQR: 291.74–1478.06; p<0.001). When combining calories from EN and propofol, AmsterdamUMCdb patients still received more daily total calories, with a median of 1727.66 kcal (IQR: 1192.90–2059.41) compared to 1307.24 kcal (IQR: 705.00–1802.50) in MIMIC-IV (p<0.001).

In contrast, MIMIC-IV patients received more daily propofol, reflected by both a higher volume (median 31.25 mL [IQR: 0.00–564.98] vs. 0.000 mL [IQR: 0.00–19.20]; p<0.001) and more propofol-derived calories (34.38 kcal [IQR: 0.00–621.48] vs. 0.00 kcal [IQR: 0.00–21.12]; p<0.001). These findings highlight differences in nutrition delivery and sedation practices driven by varying clinical practices between the AmsterdamUMCdb and MIMIC-IV institutions.

The proportion of patients with hypocaloric feeding decreased over hospital course in both datasets (Table 2). On day 3, 40.8% of patients in AmsterdamUMCdb and 53.13% in MIMIC-IV achieved hypocaloric feeding. By day 7, these proportions declined to 25.39% and 35.33%, respectively.

**Table 2.**
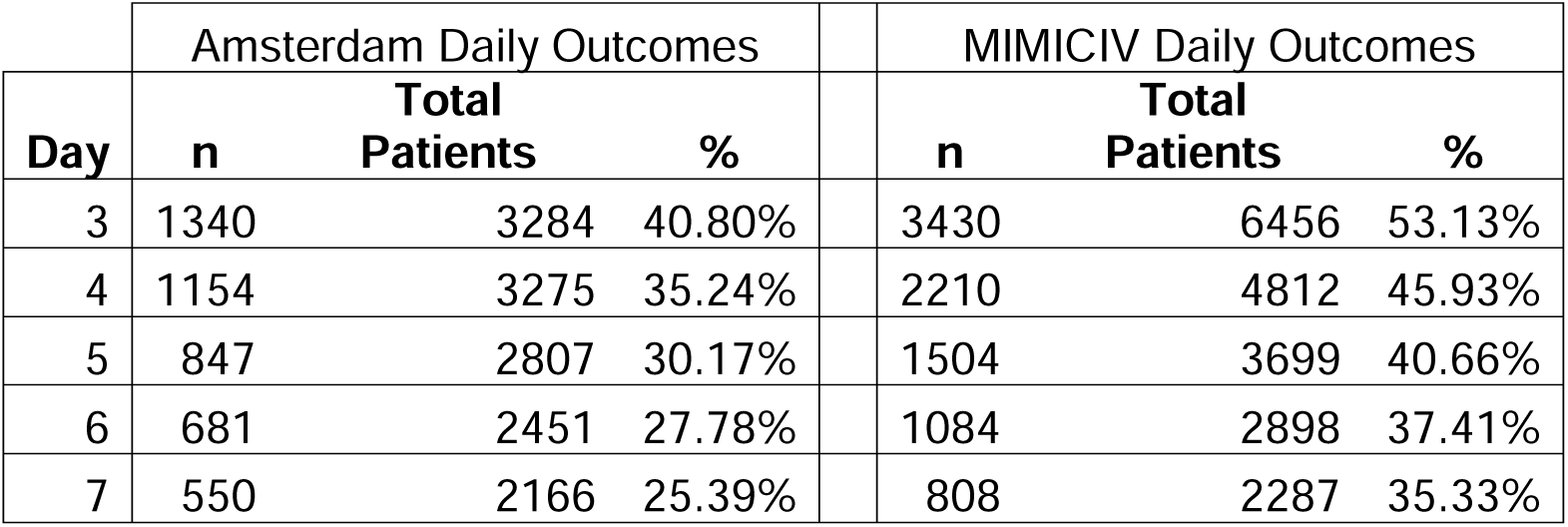
Patients Attaining Hypocaloric Feeding Over Study Timeline

| Day | Amsterdam Daily Outcomes |  |  |  | MIMICIV Daily Outcomes |  |  |
| --- | --- | --- | --- | --- | --- | --- | --- |
|  | n | Total Patients | % |  | n | Total Patients | % |
| 3 | 1340 | 3284 | 40.80% |  | 3430 | 6456 | 53.13% |
| 4 | 1154 | 3275 | 35.24% |  | 2210 | 4812 | 45.93% |
| 5 | 847 | 2807 | 30.17% |  | 1504 | 3699 | 40.66% |
| 6 | 681 | 2451 | 27.78% |  | 1084 | 2898 | 37.41% |
| 7 | 550 | 2166 | 25.39% |  | 808 | 2287 | 35.33% |

### NutriSighT Performance

The overall modeling approach of NutriSighT is shown in Figure 1. We evaluated NutriSighT’s performance across six days post-intubation (Table 3, Figure 2). On the internal test set from AmsterdamUMCdb, the Receiver Operating Characteristic Area Under the Curve (AUROC) started at 0.84 (95% CI: 0.83 - 0.84) on day 1 and was 0.73 (95% CI: 0.70 - 0.77) by day 6. External validation with the MIMIC-IV dataset revealed a similar trend, with AUROC value of 0.77 (95% CI: 0.77 - 0.78) on day 1 and 0.70 (95% CI: 0.69 - 0.71) on day 6. The model demonstrated an overall AUROC of 0.81 (95% CI: 0.81 - 0.82) on the internal test set and 0.76 (95% CI: 0.75 - 0.76) on the external validation dataset, reflecting strong discriminatory performance. The Area Under the Precision-Recall Curve (AUPRC) was 0.70 (95% CI: 0.70 - 0.72) and 0.70 (95% CI: 0.69 - 0.70) for the internal test set and external validation datasets, respectively. Additionally, the Brier score on the external validation dataset was 0.21, indicating moderate accuracy in probability predictions.

**Figure 1.**
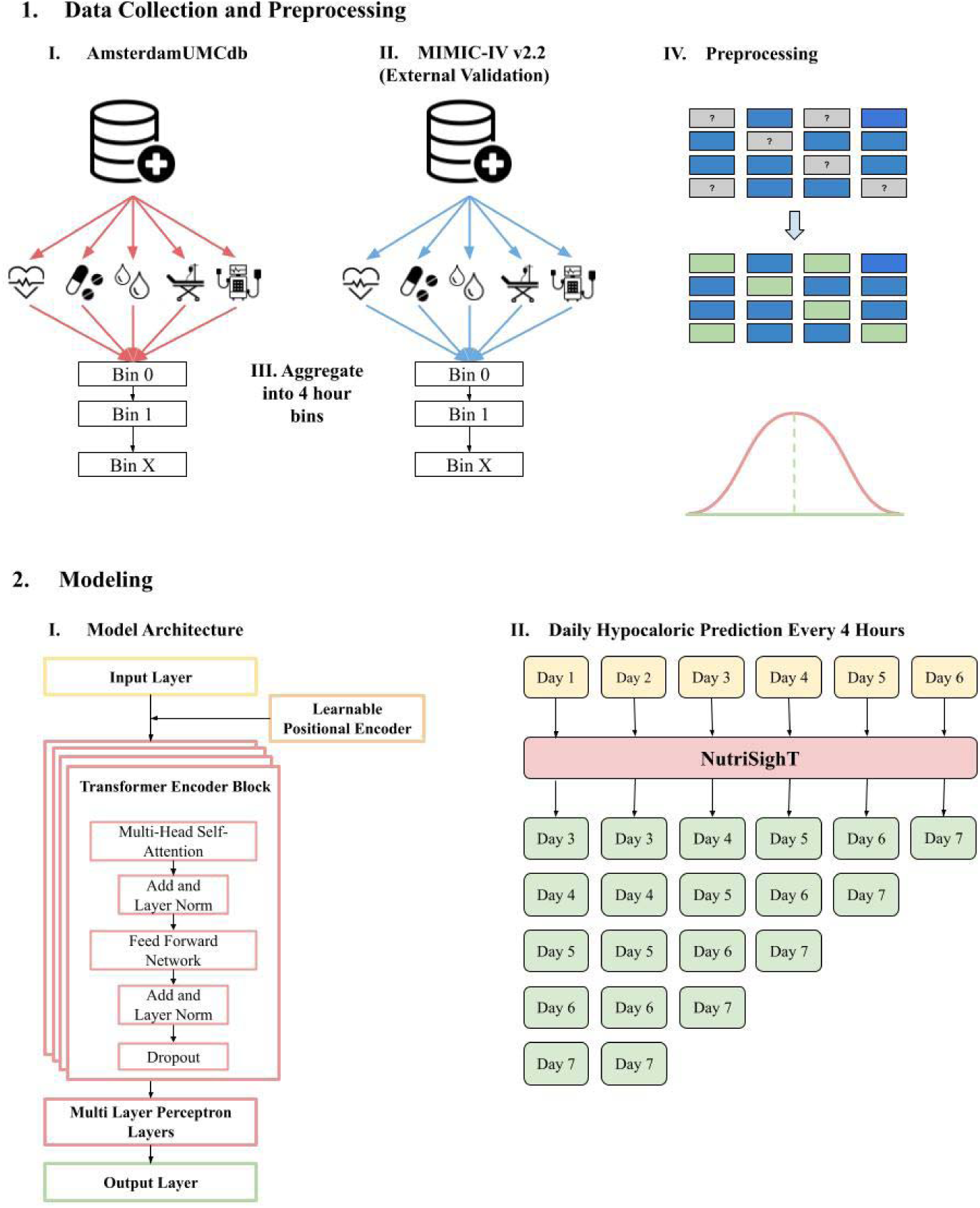
Overview of Modeling Approach

**Table 3.** Model Performance Summarized by Day

|  | *Internal Train* | *Internal Test* | *External Validation* |
| --- | --- | --- | --- |
| **AUROC (95% CI)** |  |  |  |
| **Day 1 (Timesteps 1-6)** | 0.87 (0.86, 0.87) | 0.84 (0.83, 0.84) | 0.77 (0.77, 0.78) |
| **Day 2 (Timesteps 7-12)** | 0.86 (0.86, 0.87) | 0.84 (0.82, 0.84) | 0.77 (0.77, 0.78) |
| **Day 3 (Timesteps 13-18)** | 0.88 (0.87, 0.88) | 0.82 (0.81, 0.83) | 0.76 (0.77, 0.78) |
| **Day 4 (Timesteps 19-24)** | 0.86 (0.86, 0.87) | 0.80 (0.78, 0.81) | 0.76 (0.76, 0.76) |
| **Day 5 (Timesteps 25-30)** | 0.84 (0.83, 0.84) | 0.75 (0.73, 0.77) | 0.74 (0.73, 0.74) |
| **Day 6 (Timesteps 30-36)** | 0.80 (0.78, 0.81) | 0.73 (0.70, 0.77) | 0.70 (0.69, 0.71) |
| **AUPRC (95% CI)** |  |  |  |
| **Day 1 (Timesteps 1-6)** | 0.78 (0.77, 0.78) | 0.74 (0.72, 0.76) | 0.73 (0.73, 0.73) |
| **Day 2 (Timesteps 7-12)** | 0.78 (0.78, 0.79) | 0.73 (0.71, 0.75) | 0.73 (0.73, 0.73) |
| **Day 3 (Timesteps 13-18)** | 0.77 (0.77, 0.78) | 0.71 (0.69, 0.73) | 0.71 (0.70, 0.71) |
| **Day 4 (Timesteps 19-24)** | 0.74 (0.73, 0.75) | 0.63 (0.60, 0.65) | 0.65 (0.65, 0.66) |
| **Day 5 (Timesteps 25-30)** | 0.69 (0.68, 0.70) | 0.57 (0.54, 0.61) | 0.62 (0.61, 0.63) |
| **Day 6 (Timesteps 30-36)** | 0.58 (0.56, 0.60) | 0.53 (0.48, 0.57) | 0.55 (0.54, 0.57) |

**Figure 2.**
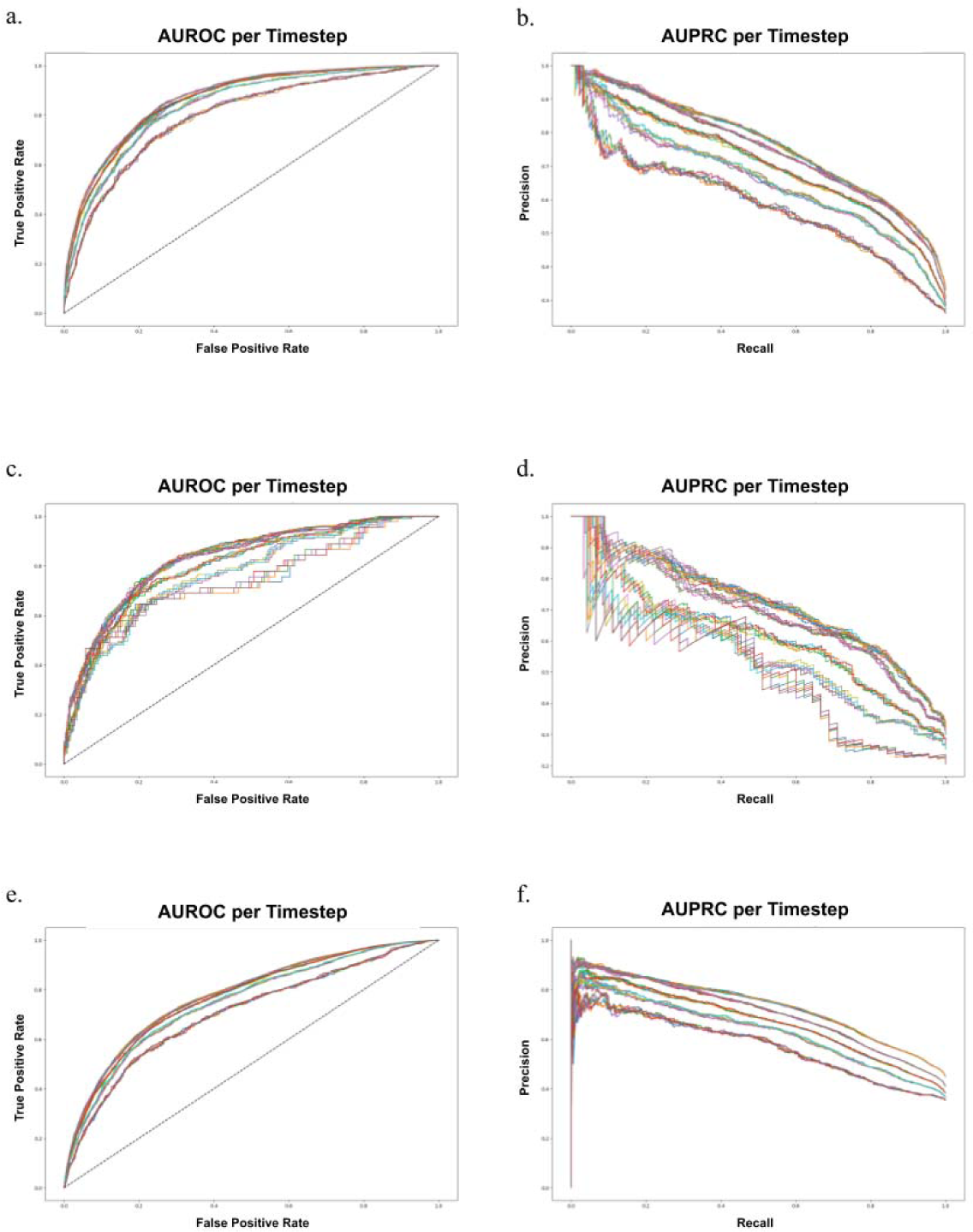
Model Performance on Internal Test and External Validation. *Plots a and b correspond to the AUROC and AUPRC on the training data that the model was developed, while plots c and d correspond to the internal test set, and plots e and f correspond to the external validation set*.

To assess the calibration of the model’s predicted probabilities more thoroughly, we generated a calibration plot (Figure 3), which compare the predicted probabilities with the observed outcomes across different probability bins. The X-axis represents the mean predicted probability of hypocaloric feeding, and the Y-axis shows the fraction of patients who actually received hypocaloric feeding within each probability bin. Figure 3 illustrates the calibration of the model on the external validation dataset. The plot shows that for probability bins below 0.5, the model’s predictions align closely with the observed outcomes, indicating good calibration in this range. However, for probability bins above 0.5, the model tends to overestimate the risk of hypocaloric feeding.

**Figure 3.**
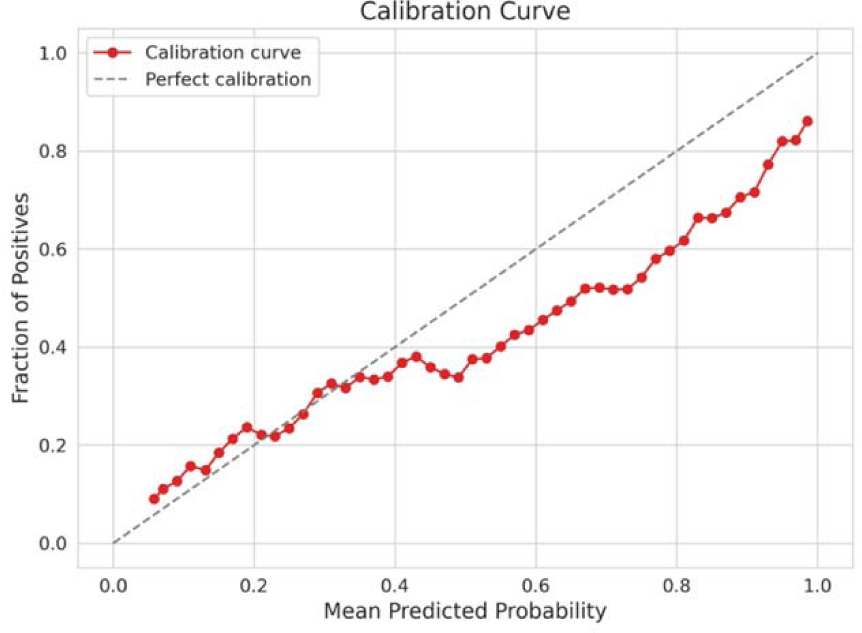
NutriSighT Calibration Curve

To determine optimal cutoff points for clinical decision-making, we further evaluated the model’s performance across various probability thresholds on the external validation data (Table 4). Sensitivity, specificity, positive predictive value (PPV), and negative predictive value (NPV) were calculated at thresholds ranging from 0.1 to 0.9. At a threshold of 0.5, the model achieved a sensitivity of 75%, specificity of 61%, PPV of 58%, and NPV of 77%. As the threshold increased, sensitivity decreased while specificity increased, illustrating the trade-off between identifying true positives and minimizing false positives. For instance, at a threshold of 0.7, sensitivity decreased to 50%, but specificity increased to 83%, and PPV improved to 69%.

**Table 4.** Model Threshold Testing

| Threshold | Sensitivity | Specificity | PPV | NPV |
| --- | --- | --- | --- | --- |
| 0.1 | 0.97 | 0.18 | 0.46 | 0.88 |
| 0.2 | 0.89 | 0.37 | 0.51 | 0.83 |
| 0.3 | 0.85 | 0.48 | 0.55 | 0.81 |
| 0.4 | 0.81 | 0.54 | 0.56 | 0.80 |
| 0.5 | 0.75 | 0.61 | 0.58 | 0.77 |
| 0.6 | 0.65 | 0.71 | 0.62 | 0.73 |
| 0.7 | 0.50 | 0.83 | 0.69 | 0.69 |
| 0.8 | 0.35 | 0.93 | 0.78 | 0.66 |
| 0.9 | 0.13 | 0.98 | 0.85 | 0.61 |

### Feature Importance

We conducted permutation importance analysis by shuffling each feature in the validation dataset and measuring the resulting change in binary cross-entropy loss (Figure 4). The PAO /FIO ratio (0.08) and indirect bilirubin (0.07) were identified as the most influential positive predictors for hypocaloric feeding, indicating that higher values of these features increase the model’s likelihood of predicting hypocaloric feeding. Conversely, anion gap (−0.06), LDH (−0.06), and pH (−0.05) were significant negative predictors, meaning that higher values of these features decrease the model’s likelihood of predicting hypocaloric feeding.

**Figure 4.**
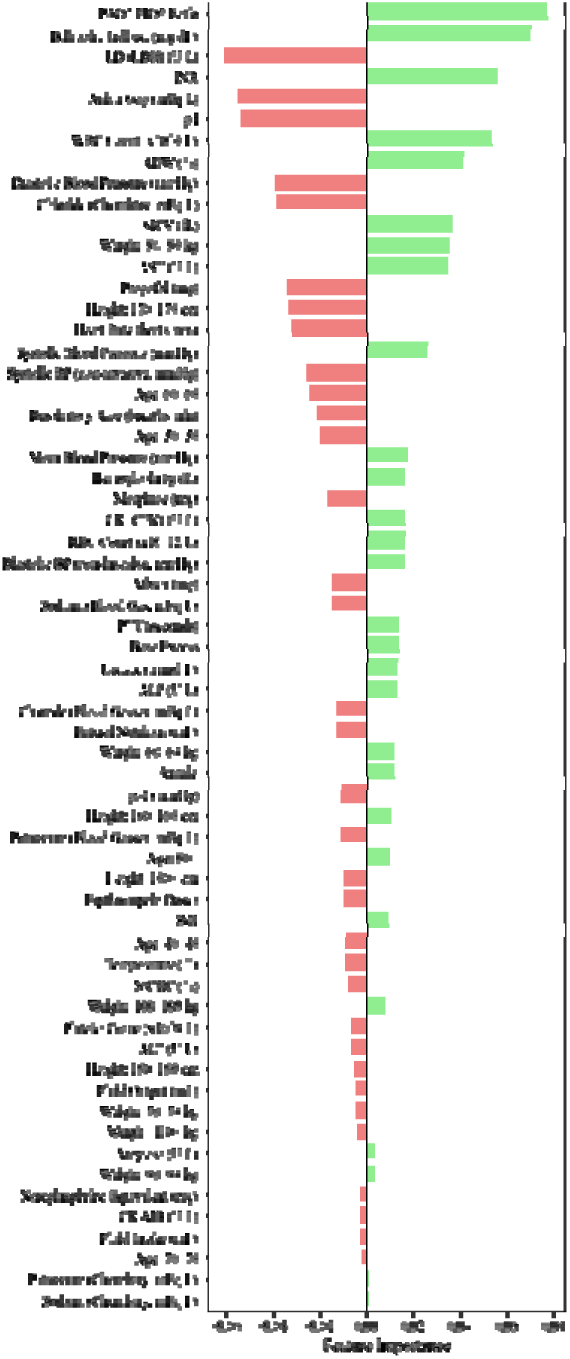
Feature Importances for NutriSighT. *Positive values (green) indicate that the feature increases the likelihood of hypocaloric feeding, while negative values (red) indicate that the feature decreases the likelihood*.

## DISCUSSION

We have developed and externally validated NutriSighT, an interpretable AI model using a novel transformer architecture to identify critically ill patients at risk of receiving hypocaloric enteral nutrition in the late acute phase of their illness. The model was trained on AmsterdamUMCdb, a European dataset, and validated on MIMIC-IV, a US dataset, demonstrating its ability to generalize across diverse patient populations [15–18]. NutriSighT exhibited strong discriminatory performance, with robust AUROC scores across both internal and external datasets, indicating its ability to differentiate between patients at higher and lower risk accurately. Calibration analysis revealed good alignment between predicted probabilities and observed outcomes.

The time-series nature of the data makes transformer architecture well-suited for capturing temporal dependencies and complex patterns [14]. This approach is ideal for predicting dynamic outcomes, such as hypocaloric enteral nutrition in critically ill patients. By modeling these temporal dynamics, NutriSighT can deliver timely and accurate predictions that adapt to changes in patient status. NutriSighT generates predictions every 4 hours, providing clinicians with actionable time windows to adjust treatment plans as necessary. The model exhibited strong discriminative ability, as demonstrated by its AUC values, and its AUPRC and calibration further support its potential for clinical integration. These features enable NutriSighT to reliably identify patients at risk for hypocaloric feeding and guide timely interventions.

Feature importance analysis revealed key predictors influencing the model’s decisions, offering insights into factors associated with hypocaloric enteral nutrition. In this case, the PAO2/FIO2 ratio and indirect bilirubin levels were identified as strong predictors for risk for hypocaloric nutrition. These findings reflect the model’s focus on clinically relevant factors, providing a glimpse into its decision-making process. It also highlights the model’s potential to capture meaningful patterns in the data, which could support personalized nutrition strategies in the ICU.

Despite the critical role of enteral nutrition in mechanically ventilated patients, optimizing feeding strategies remains a challenge [13]. Prior studies have yielded inconsistent results on the impact of different feeding strategies in these patients, highlighting the complexity of optimizing nutritional interventions. For example, an observational study of over 2,700 mechanically ventilated patients in 167 ICUs where most patients received hypocaloric nutrition, found that an increase of 1000 kcal/day was associated with lower 60-day mortality and increased number of ventilator free days [19]. Lower mortality [8] [20] [21] and shorter duration of mechanical ventilation [6] with isocaloric nutrition has also been shown in other studies.

Conversely, other studies have found either no differences in ventilator-free days, mortality, or infectious complications between the two nutritional strategies [22], or found longer time to readiness for ICU discharge among patients receiving isocaloric nutrition [23]. These conflicting results likely reflect the heterogeneity of critically ill patients, highlighting the challenges of applying a one-size-fits-all approach to nutrition.

These discrepancies underscore the need for personalized nutrition approaches, particularly in the late acute phase of critical illness, when patient conditions and metabolic needs evolve rapidly. NutriSighT addresses this need by dynamically identifying patients who are likely to receive hypocaloric nutrition in the late acute phase. By providing a precise identification of these patients, NutriSighT can guide more personalized interventions and enrich clinical trials that explore the efficacy of tailored nutritional regimens. Furthermore, NutriSighT lays the groundwork for further research into barriers to implementation and strategies to personalize nutrition in this vulnerable patient population.

This study has some limitations. First, as a retrospective analysis, it is subject to inherent biases, including selection bias and potential confounding factors, which may affect the interpretation of results. However, it showed good generalizability as supported by its strong performance across both internal and external validation datasets. Notably, the Brier score on the external validation set was 0.21 suggesting that its predicted likelihoods are reasonably well-calibrated to the actual outcomes. Second, like many studies in this field, caloric requirements are estimated using guideline recommendations rather than indirect calorimetry, as latter were unavailable. While this could introduce inaccuracies, it reflects real-world clinical practice where indirect calorimetry is rarely used, thereby enhancing the generalizability of our findings. Future prospective studies incorporating direct calorimetry data may provide deeper insights and further validate the model’s utility. Third, although this study focuses on dynamically identifying patients likely to receive hypocaloric nutrition, addressing the broader spectrum of nutritional risk including underfeeding, overfeeding, or interruptions in feeding requires further exploration. This work represents an important step forward by paving the way for more nuanced approaches to personalized nutritional strategies in critically ill patients. Finally, the observed decline in model performance from day 1 to day 6 may be partly due to the decreasing sample size, as fewer patients remain on mechanical ventilation over time. With a smaller number of patients later in the ventilation period, the model has fewer data points to make predictions, which can lead to a slight decrease in performance. Despite this, the model still demonstrates strong performance overall, highlighting its ability to identify at-risk patients early and throughout the ventilation period. These results highlight the model’s robustness and potential for deployment across diverse patient populations and healthcare settings.

In conclusion, we developed NutriSighT, an interpretable transformer model designed to identify mechanically ventilated, critically ill patients likely to only receive hypocaloric enteral nutrition. NutriSighT has the potential to facilitate timely nutritional interventions in critically ill patients. Future studies should focus on integrating this model into trial designs to identify high-risk patients and optimize their nutritional strategies.

## METHODS

### Data Sources

In this retrospective study we utilized data from two independent ICU datasets-the Amsterdam University Medical Centers Database (AmsterdamUMCdb) and the Medical Information Mart for Intensive Care IV (MIMIC-IV v2.2) (Figure 4).

AmsterdamUMCdb is a highly granular ICU dataset from the European Union, containing deidentified electronic health records of ICU patients from the Amsterdam University Medical Centers in the Netherlands [16]. It includes admission data spanning 2003 to 2016 and encompasses approximately 1 billion data points including demographics, vital signs, laboratory tests and medications from over 20,000 ICU admissions. In contrast, MIMIC-IV is a United States based, single-center, de-identified database comprising electronic health records data from over 70,000 ICU admissions at the Beth Israel Deaconess Medical Center, with ICU admission data ranging from 2008 to 2019 [17,18].

### Study Population

We included patients 18 years or older who were mechanically ventilated in the ICU for at least 72 hours. Patients receiving total parenteral nutrition or peripheral parenteral nutrition during the ventilation event were excluded. We also excluded patients with missing height or weight data, or ambiguous data regarding tube feeds that did not allow us to calculate the amount of tube feeds administered (Supplementary Figure 1).

### Outcomes

The primary outcome of this study was to identify patients likely to receive hypocaloric enteral nutrition on a given day between days 3-7 of mechanical ventilation among ICU patients. Predictions were censored to the day prior if a patient was extubated, died, or transferred out of the ICU.

Consistent with the recommendations of the American Society for Parenteral and Enteral Nutrition (ASPEN) guidelines [11], we estimated caloric requirements using weight-based equations adjusted according to the patient’s Body Mass Index (BMI) as below:

⍰ **For patients with BMI < 30 kg/m²:** 25 kcal per kilogram of actual body weight per day.

⍰ **For patients with 30 BMI 50 kg/m²:** 11 kcal per kilogram of actual body weight per day.

⍰ **For patients with BMI > 50 kg/m²:** 22 kcal per kilogram of adjusted body weight per day.

For BMI > 50 kg/m², adjusted body weight (kilogram) was calculated as [24,25]:

⍰ **Females:** 45.36 + 2.27 × (Height (cm) - 152.4)

⍰ **Males:** 48.08 + 2.72 × (Height (cm) - 152.4)

Hypocaloric enteral nutrition was defined as receipt of less than 70% of the calculated caloric requirements on a given day [13,26], with caloric intake determined by a combination of enteral nutrition and propofol.

### Feature Extraction

We extracted a comprehensive set of features from the AmsterdamUMCdb and MIMIC-IV databases to capture the clinical characteristics of ICU patients. The data was collected starting from the time of ICU admission or the time of intubation, if the latter occurred after ICU admission. It continued for up to 7 days after the start of mechanical ventilation, with earlier censoring in the case of extubation, death, or transfer out of the ICU. The features included demographics, vital signs, laboratory results, medications administered, enteral nutrition, fluid intake, fluid output and enteral nutrition. Demographics included age, sex, height, weight, and body mass index (BMI). Vital signs included heart rate, systolic and diastolic blood pressures, mean arterial pressure, respiratory rate, and temperature. Laboratory results included oxygen saturation and PaO /FiO ratio, pH, base excess, lactate, sodium, potassium, chloride, anion gap, hemoglobin, hematocrit, mean corpuscular volume, mean corpuscular hemoglobin concentration, red blood cell count, white blood cell count, platelet count, red cell distribution width, international normalized ratio, partial thromboplastin time, alanine aminotransferase, aspartate aminotransferase, alkaline phosphatase, indirect bilirubin, lactate dehydrogenase, amylase, creatine kinase, CK-MB, blood urea nitrogen and creatinine. We also included data regarding medications administered such as vasopressors (in norepinephrine equivalent doses), sedatives and analgesics (lorazepam, morphine, propofol), and prokinetics (erythromycin and metoclopramide) [27]. We further extracted the amount of enteral nutrition administered and calories delivered by enteral nutrition and propofol.

### Data Pre-processing

To capture the temporal changes during each patient’s ICU stay, we structured the data into 4-hour time intervals, starting from the time of ICU admission or the time of intubation, if the latter was after ICU admission and ending at the earlier of extubation or 7 days after intubation. Clinical variables were summed or averaged within each time interval as appropriate. We excluded features with more than 40% missingness to ensure data quality and reliability, following standard practices in data analysis [28]. Outliers were identified and excluded based on clinical expertise, removing data points that were physiologically implausible or indicative of measurement errors.

Consistent with standard methods for handling missing data in these datasets, we used forward fill imputation for all features (except for medications administered, fluid intake, and enteral nutrition which were treated as zero when missing) and applied k-nearest neighbor (k-NN) imputation (k=5) to fill in any remaining missing values [29–31].

As AmsterdamUMCdb provides age, height, and weight in pre-defined subgroups (Table 1), we applied a similar approach in MIMIC-IV and encoded them using one-hot encoding to transform them into a binary format suitable for the model. Continuous variables were standardized using z-score normalization to ensure that all features contributed equally to the model training and to facilitate the convergence of the optimization algorithm. This step transformed the variables to have a mean of zero and a standard deviation of one, reducing the risk of features with larger numerical ranges dominating the learning process.

### Model Development

At the core of NutriSighT’s functionality is its capability of making predictions at every 4-hour interval. At each timestep, the model updates its assessment based on the most recent patient data and predicts the hypocaloric feeding status separately for Days 3, 4, 5, 6, and 7. These predictions are censored up to the day before the earliest occurrence of extubation, death, or discharge from the ICU, ensuring that the predictions remain relevant to the patient’s current clinical trajectory.

The model input comprises sequential data structured into 4-hour intervals, spanning up to seven days of mechanical ventilation. Each input sequence has a shape 36 x 62 (the number of 4-hour time bins over 6 prediction days by the number of clinical features). Traditional transformer models use fixed sinusoidal positional encodings to incorporate the order of input sequences [14]. However, we implemented a learnable positional encoder, which allows the model to learn optimal positional representations during training [32]. This approach adds trainable positional embeddings to the input sequences, enabling the model to better capture temporal dynamics and improve performance on sequential tasks.

NutriSighT is comprised of four stacked Transformer Encoder Blocks, each featuring multi-head self-attention mechanisms with four heads and a head size of 512. This architecture handles sequential data by capturing long-range dependencies through self-attention, enhancing the model’s ability to discern complex temporal patterns inherent in clinical data [14,15]. Following the self-attention layers, dropout layer normalization are applied. Following the transformer encoder layers, the model integrates a series of Multi-Layer Perceptron (MLP) Layers with 312, 64, and 48 units, respectively. Each dense layer is accompanied by dropout (35%) and L2 regularization (10^-5^). The final output layer employs a sigmoid activation function, generating probabilistic predictions for each outcome day.

These strategies were employed to prevent overfitting and optimize convergence, ensuring that NutriSighT effectively generalizes to unseen data. The combination of transformer architecture, learnable positional encodings, and robust regularization techniques enables NutriSighT to deliver accurate and timely predictions, thereby facilitating personalized nutritional interventions in the ICU setting.

### Rolling Prediction Framework

The prediction schedule followed a dynamic, rolling approach as below (Figure 5):

- **Days 1 and 2**: Predicted hypocaloric feeding status for each day from **Day 3 to Day 7**, with prediction updated every 4 hours.
- **Day 3**: Predicted for each day from **Day 4 to Day 7**, updated every 4 hours.
- **Day 4**: Predicted for each day from **Day 5 to Day 7**, updated every 4 hours.
- **Day 5**: Predicted for **Days 6 and 7**, updated every 4 hours.
- **Day 6**: Predicted for **Day 7**, updated every 4 hours.

**Figure 5.**
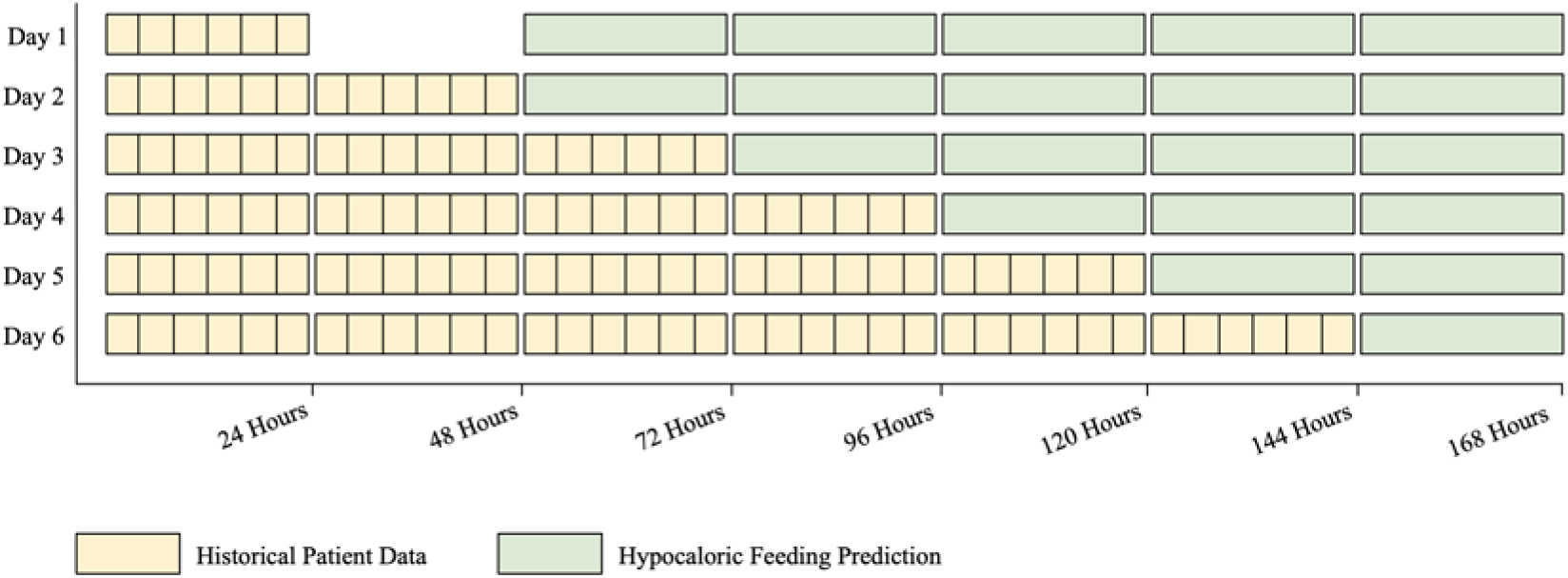
Model Rolling Prediction Schema

This approach ensured continuous updates, allowing clinicians to adapt nutritional strategies proactively. By predicting multiple days ahead at each timestep, the model supports better planning and timely interventions.

### Training and Validation

We split the AmsterdamUMCdb dataset into a training set (80%), an internal validation set (10%), and an internal test set (10%). The internal validation set was used during training to prevent overfitting and tune hyperparameters. To address class imbalance, we employed class weighting, a technique that adjusts the contribution of each class to the loss function during training [33]. Specifically, we calculated class weights inversely proportional to the frequency of positive and negative outcomes, assigning higher weights to the minority class and lower weights to the majority class. By modifying the loss function in this way, the model was encouraged to pay greater attention to underrepresented outcomes. This approach reduced the risk of the model disproportionately favoring the majority class and thus helped the model learn patterns associated with both outcomes more effectively.

We trained the model using the Adam optimizer with a learning rate of 5×10 and applied early stopping and learning rate reduction callbacks to prevent overfitting and optimize training time. To ensure that the performance was clinically meaningful, we evaluated the model on the internal test set and external validation set using metrics sensitive to class imbalance, such as precision and recall. These metrics reflect the model’s ability to not only classify patients accurately but also to reliably identify those at highest risk, thereby enhancing the real-world utility of our predictive framework.

### Statistical Analysis

We assessed model performance using several statistical metrics to evaluate its predictive accuracy and generalizability. The Receiver Operating Characteristic Area Under the Curve (AUROC) was used to evaluate the model’s ability to discriminate between patients who would and would not receive hypocaloric enteral nutrition. The Area Under the Precision-Recall Curve (AUPRC) was calculated to assess the trade-off between precision and recall. We also analyzed feature importance using a permutation-based method, which allowed us to evaluate the contribution of each feature to NutriSighT’s predictive performance.

To evaluate the calibration of the model’s predicted probabilities, we computed the Brier score and generated calibration plots, which visually assess how closely predicted probabilities align with actual outcomes across probability bins. Comparative statistical analyses were conducted using the Mann-Whitney U test for continuous variables and the chi-squared test for categorical variables. The trained model was externally validated on the MIMIC-IV dataset to assess its generalizability across different patient populations and clinical settings. This external validation ensured that the model’s performance is robust and applicable to diverse ICU environments.

## Data Availability

Publicly available datasets were analyzed in this study. The dataset used in this study, MIMIC-IV, is available at https://mimic.physionet.org/, and the AmsterdamUMCdb dataset is available at https://amsterdammedicaldatascience.nl/amsterdamumcdb/.

## Acknowledgement

None

## Funding

This study was supported by the National Institutes of Health (NIH) grant K08DK131286 awarded to Ankit Sakhuja. The funder had no role in study design, data collection and analysis, decision to publish, or preparation of the manuscript.

## Competing Interests

GNN is a founder of Renalytix, Pensieve, Verici and provides consultancy services to AstraZeneca, Reata, Renalytix, Siemens Healthineer and Variant Bio, serves a scientific advisory board member for Renalytix and Pensieve. He also has equity in Renalytix, Pensieve and Verici. LC is a consultant for Vifor Pharma INC and has received honorarium from Fresenius Medical Care. All remaining authors have declared no conflicts of interest.

## Notes

### Funding Statement

This work was supported by the National Institutes of Health (NIH) grants K08DK131286 awarded to Ankit Sakhuja. The content is solely the responsibility of the authors and does not necessarily represent the official views of the National Institutes of Health.

### Author Declarations

Study used publicly available datasets - AmsterdamUMCdb available at https://amsterdammedicaldatascience.nl; and MIMIC-IV available at https://physionet.org/content/mimiciv/3.1/

