## Supplementary for "NutriSighT: Interpretable Transformer Model for Dynamic Prediction of Hypocaloric Enteral Nutrition in Mechanically Ventilated Patients"

**Supplementary Table 1. Additional baseline characteristics**

| | | AmsterdamUMCdb | MIMIC-IV | p-value |
|---|---|---|---|---|
| **Medication Dosage** | | | | |
| | *Ativan (mg), median (IQR)* | 0 (0, 0) | 0 (0,0) | <0.001 |
| | *Morphine (mg), median (IQR)* | 0 (0, 0) | 0 (0,0) | 0.141 |
| | *Propofol (mg), median (IQR)* | 0 (0, 119.5) | 0 (0, 411.45) | <0.001 |
| | *Norepniephrine Equivalent (mg), median (IQR)* | 0 (0, 0.11) | 0 (0, 0.07) | <0.001 |
| | *Erythromycin Doses, median (IQR)* | 0 (0, 0) | 0 (0,0) | <0.001 |
| **Nutrition** | | | | |
| | *Enteral Nutrition (ml), median (IQR)* | 1440.00 (1000.40, 1723.80) | 756.48 (228.72, 1199.97) | <0.001 |
| | *Propofol (ml), median (IQR)* | 0.00 (0.00, 19.20) | 31.25 (0.00, 564.98) | <0.001 |
| | *Enteral Nutrition and Propofol (calories), median (IQR)* | 1727.66 (1192.90, 2059.41) | 1307.24 (705.00, 1802.50) | <0.001 |
| | *Enteral Nutrition (calories), median (IQR)* | 1702.99 (1155.26, 2039.66) | 989.20 (291.74, 1478.06) | <0.001 |
| | *Propofol (calories), median (IQR)* | 0.00 (0.00, 21.12) | 34.38 (0.00, 621.478) | <0.001 |
| **Fluids** | | | | |
| | *Fluid Intake (ml), median (IQR)* | 518.87 (387.71, 716.89) | 444.28 (266.46, 720.47) | <0.001 |

| | | | | |
|---|---|---|---|---|
| | *Fluid Output (ml), median (IQR)* | 322.42 (181.50, 548.83) | 260.00 (125.00, 483.74) | <0.001 |
| **Vital Signs** | | | | |
| | *Heart Rate (beats/min), median (IQR)* | 84.75 (72.5, 98) | 84.714 (73.4, 97) | <0.001 |
| | *Respiratory Rate (breaths /min), median (IQR)* | 18.5 (14, 23.6) | 20 (16.8, 23.8) | <0.001 |
| | *Systolic Blood Pressure (mmHg), median (IQR)* | 128.25 (113.25, 146.333) | 116.2 (105.2, 130.6) | <0.001 |
| | *Diastolic Blood Pressure (mmHg), median (IQR)* | 61.004 (54.404, 69) | 59 (52.333, 67) | <0.001 |
| | *Mean Blood Pressure (mmHg), median (IQR)* | 83 (74.5, 93.25) | 76 (69.25, 85) | <0.001 |
| | *Systolic Blood Pressure (non-invasive, mmHg), median (IQR)* | 105.4 (79.7, 126.667) | 115 (102, 130.5) | <0.001 |
| | *Diastolic Blood Pressure (non-invasive, mmHg), median (IQR)* | 60.425 (47.2, 72) | 61.857 (53.75, 71.167) | <0.001 |
| | *Temperature (℃), median (IQR)* | 36.6 (35.9, 37.2) | 37.17 (36.78, 37.61) | <0.001 |
| **Laboratory Results** | | | | |
| | *SpO2 (mmHg), median (IQR)* | 93 (78, 113) | 101 (77, 132) | <0.001 |

| | | | | |
|---|---|---|---|---|
| | *PAO2/FIO2 Ratio, median (IQR)* | 94.949 (81.25, 116.162) | 226.667 (154, 312.5) | <0.001 |
| | *pH, median (IQR)* | 7.41 (7.36, 7.45) | 7.41 (7.36, 7.45) | <0.001 |
| | *Base Excess, median (IQR)* | 2.9 (0.9, 5.4) | 0 (-2, 4) | <0.001 |
| | *Hemoglobin (g/dL), median (IQR)* | 6.5 (5.8, 7.2) | 10.6 (9.68, 11.5) | <0.001 |
| | *Chloride (Bloog Gasses, mEq/L), median (IQR)* | 106.5 (103.8, 109) | 106 (103.4, 109) | <0.001 |
| | *Potassium (Blood Gasses, mEq/L), median (IQR)* | 4.1 (3.8, 4.35) | 4.1 (3.77, 4.46) | <0.001 |
| | *Sodium (Blood Gas, mEq/L), median (IQR)* | 140 (137, 143) | 137.8 (135.2, 140) | <0.001 |
| | *Lactate (mmol/L), median (IQR)* | 1.45 (1, 2.2) | 1.4 (1.1, 2) | <0.001 |
| | *Anion Gap (mEq/L), median (IQR)* | 8.6 (6.7, 10.56) | 14 (11, 16) | <0.001 |
| | *Chloride (Chemistry, mEq/L), median (IQR)* | 107.3 (104, 111.5) | 104 (100, 109) | <0.001 |
| | *Sodium (Chemistry, mEq/L), median (IQR)* | 142 (139, 146) | 140 (137, 144) | <0.001 |
| | *Potassium (Chemistry, mEq/L), median (IQR)* | 4.1 (3.9, 4.4) | 4 (3.7, 4.4) | <0.001 |
| | *MCHC (%), median (IQR)* | 21.2 (21, 21.6) | 32.7 (31.5, 33.8) | <0.001 |

| | | | | |
|---|---|---|---|---|
| | *MCV (fL), median (IQR)* | 90.4 (88, 92.8) | 92 (88, 96) | <0.001 |
| | *Platelet Count (x10^9/L), median (IQR)* | 179 (117, 258) | 179 (110, 259) | <0.001 |
| | *RBC Count (x10^12/L), median (IQR)* | 3.6 (3.3, 3.98) | 3.12 (2.76, 3.54) | <0.001 |
| | *RDW (%), median (IQR)* | 13.15 (13.15, 13.15) | 15.6 (14.3, 17.3) | <0.001 |
| | *WBC Count (x10^9/L), median (IQR)* | 12 (8.9, 16.2) | 11.5 (8.3, 15.9) | <0.001 |
| | *INR, median (IQR)* | 1.24 (1.12, 1.4) | 1.3 (1.1, 1.5) | <0.001 |
| | *PTT (seconds), median (IQR)* | 47 (40, 59.5) | 32.8 (28.3, 43.8) | <0.001 |
| | *ALT (U/L), median (IQR)* | 41.8 (22, 89) | 39 (20, 95.6) | <0.001 |
| | *ALP (U/L), median (IQR)* | 81 (59, 120) | 98 (69, 147.45) | <0.001 |
| | *AST (U/L), median (IQR)* | 51 (29, 110) | 58 (30.4, 135) | <0.001 |
| | *Amylase (U/L), median (IQR)* | 82 (47, 143.8) | 95.6 (63, 154) | <0.001 |
| | *Bilirubin, Indirect (mg/dL), median (IQR)* | 0.482 (0.396, 0.56) | 1.82 (1.1, 2.6) | <0.001 |
| | *CK (CPK) (U/L), median (IQR)* | 249 (80, 711) | 403.2 (147, 1037) | <0.001 |
| | *CK-MB (U/L), median (IQR)* | 12.8 (6.6, 27.1) | 8.8 (4.4, 20.2) | <0.001 |
| | *LD (LDH) (U/L), median (IQR)* | 335 (232, 519) | 376.4 (271, 569.6) | <0.001 |
| **Demographics** | | | | |
| | *BMI, median (IQR)* | 24.84 (23.88, 27.76) | 28.1 (23.98, 33.66) | <0.001 |
| ***Sex*** | | | | <0.001 |

| | | | | |
|---|---|---|---|---|
| | *Female, n (%)* | 1183 (36.02) | 2661 (41.22%) | |
| | *Male, n (%)* | 2121 (64.58) | 3795 (58.78) | |
| ***Age (years)*** | | | | <0.001 |
| | *18-39* | 366 (11.15) | 591 (9.15) | |
| | *40-49* | 317 (9.65) | 577 (8.94) | |
| | *50-59* | 583 (17.75) | 1161 (17.98) | |
| | *60-69* | 801 (24.39) | 1574 (24.38) | |
| | *70-79* | 848 (25.82) | 1448 (22.43) | |
| | *80+* | 369 (11.24) | 1105 (17.12) | |
| ***Height (cm)*** | | | | <0.001 |
| | *0-159* | 149 (4.54) | 1198 (18.56) | |
| | *160-169* | 815 (24.82) | 1908 (29.55) | |
| | *170-179* | 1220 (37.15) | 2213 (34.28) | |
| | *180-189* | 925 (28.17) | 1029 (15.94) | |
| | *190+* | 175 (5.33) | 108 (1.67) | |
| ***Weight (kg)*** | | | | <0.001 |
| | *0-59* | 280 (8.53) | 856 (13.26) | |
| | *60-69* | 563 (17.14) | 1030 (15.95) | |
| | *70-79* | 888 (27.04) | 1156 (17.91) | |
| | *80-89* | 856 (26.07) | 1030 (15.95) | |
| | *90-99* | 416 (12.67) | 834 (12.92) | |
| | *100-109* | 138 (4.20) | 617 (9.56) | |
| | *110+* | 143 (4.35) | 933 (14.45) | |

*Baseline characteristics, including medication dosages, nutrition, fluids, vital signs, labs, and demographics, are reported as median (IQR) or n (%). Data are aggregated over four 4-hour periods, except nutrition, which is over 24 hours.*

**Supplementary Figure 1. Study Cohort**

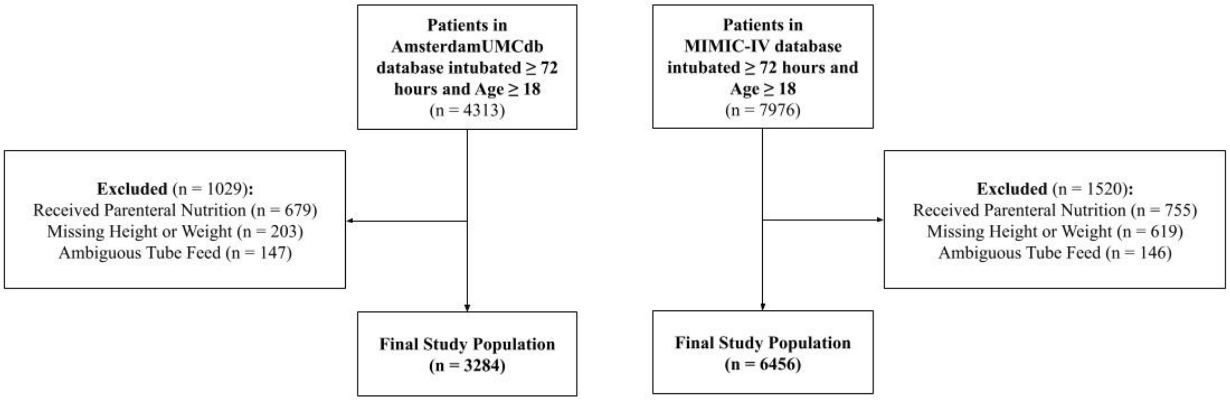


*This diagram outlines patient inclusion and exclusion criteria. Patients intubated for ≥72 hours and aged ≥18 were included, with exclusions for parenteral nutrition, missing height or weight, and ambiguous tube feed documentation, resulting in final study populations of 3,284 (AmsterdamUMCdb) and 6,456 (MIMIC-IV).*
